# Before the Surge: Molecular Evidence of SARS-CoV-2 in New York City Prior to the First Report

**DOI:** 10.1101/2021.02.08.21251303

**Authors:** Matthew M. Hernandez, Ana S. Gonzalez-Reiche, Hala Alshammary, Shelcie Fabre, Zenab Khan, Adriana van De Guchte, Ajay Obla, Ethan Ellis, Mitchell J. Sullivan, Jessica Tan, Bremy Alburquerque, Juan Soto, Ching-Yi Wang, Shwetha Hara Sridhar, Ying-Chih Wang, Melissa Smith, Robert Sebra, Alberto E. Paniz-Mondolfi, Melissa R. Gitman, Michael D. Nowak, Carlos Cordon-Cardo, Marta Luksza, Florian Krammer, Harm van Bakel, Viviana Simon, Emilia Mia Sordillo

**Author notes:** Correspondence to (V.S.), (H.B.), (E.M.S.). Contributed equally (Matthew M. Hernandez, Ana S. Gonzalez-Reiche). Co-senior authors (Harm van Bakel, Viviana Simon, Emilia Mia Sordillo).

## Abstract

New York City (NYC) emerged as a coronavirus disease 2019 (COVID-19) epicenter in March 2020, but there is limited information regarding potentially unrecognized severe acute respiratory syndrome coronavirus 2 (SARS-CoV-2) infections before the first reported case. We utilized a sample pooling strategy to screen for SARS-CoV-2 RNA in de-identified, respiratory pathogen-negative nasopharyngeal specimens from 3,040 patients across our NYC health system who were evaluated for respiratory symptoms or influenza-like illness during the first 10 weeks of 2020. We obtained complete SARS-CoV-2 genome sequences from samples collected between late February and early March. Additionally, we detected SARS-CoV-2 RNA in pooled specimens collected in the week ending 25 January 2020, indicating that SARS-CoV-2 caused sporadic infections in NYC a full month before the first officially documented case.

**ONE SENTENCE SUMMARY:** Molecular surveillance demonstrates that SARS-CoV-2 caused influenza-like illness in NYC before the first reported case.

## MAIN TEXT

The first cases of coronavirus disease 19 (COVID-19), caused by severe acute respiratory syndrome coronavirus 2 (SARS-CoV-2), were observed in China in December 2019 (*1, 2*). Within weeks, cases were reported in other countries in Asia, as well as in Europe and North America. In the United States (US), the first SARS-CoV-2 infection was confirmed by the US Centers for Disease Control and Prevention (CDC) on 20 January 2020 (*3*). During the following weeks, sporadic cases were reported throughout the US. When the first case in New York State (NYS) was diagnosed in New York City (NYC) on 29 February 2020 (*4*), the NYC metropolitan area quickly emerged as an early epicenter of the pandemic.

We previously documented multiple independent introductions of SARS-CoV-2 into the NYC metropolitan area based on SARS-CoV-2 genomes obtained from 84 patients with COVID-19 receiving care at acute care hospitals and affiliated outpatient facilities of the Mount Sinai Health System (MSHS) during March 2020 (*4*). Based on phylogenetic reconstructions, we estimated that these independent SARS-CoV-2 introductions occurred early in February 2020 (*4, 5*); this timeframe is further supported by our recent cross-sectional serosurvey of MSHS patients (*6*). However, prior to mid-March, 2020, COVID-19 case detection was limited by restricted availability of diagnostic testing and overlap in symptom presentation with other respiratory and viral illnesses. Thus, direct molecular evidence of SARS-CoV-2 in NYC prior to the first reported case is lacking.

To systematically delineate the arrival of SARS-CoV-2 in NYC, we secured 3,040 residual nasopharyngeal swab specimens collected in viral transport medium that were banked from patients with respiratory symptoms or influenza-like illness who presented to the MSHS during the first 10 weeks of 2020 (epidemiological weeks ending on 4 January to 7 March), but were found negative by diagnostic molecular amplification testing for routine respiratory pathogens. The number of these residual respiratory pathogen-negative (RPN) specimens collected at each MSHS site varied among the MSHS hospitals as well as from week to week (**Fig. S1A**).

To increase our screening capacity and ensure specimen de-identification, we combined equal volumes of viral transport media from 10 distinct RPN specimens into single tubes, yielding 304 pools which underwent nucleic-acid amplification testing (NAAT) for SARS-CoV-2 (**Fig. 1A**) using the Roche Diagnostics cobas® 6800 SARS-CoV-2 Test. This assay, which has emergency use authorization from the US Food and Drug Administration for the detection of SARS-CoV-2 in clinical specimens, evaluates samples for the presence of the SARS-CoV-2-specific ORF1ab gene (target 1, T1) and the pan-*Sarbecovirus* envelope (E)-gene (target 2, T2). Of the 304 RPN pools, 9 (3%) tested positive (both targets or only T1 detected), 8 (2.6%) tested presumptive positive (only T2 detected), and 287 (94.4%) were negative (neither target detected) (**Fig. 1B**). Five presumptive positive RPN pools contained specimens from patients treated at two distinct MSHS hospitals (A and C), collected during the weeks ending on 18 January, 25 January, and 1 February (**Fig. 1C**). None of the RPN pools comprised of specimens collected during the following three weeks yielded detectable SARS-CoV-2 RNA. However, for specimens collected in the week ending 29 February, SARS-CoV-2 RNA was detected in 5.4% of RPN pools (3.6% positive, 1.8% presumptive positive); this percentage increased to 33.3% (25.9% positive, 7.4% presumptive positive) for RPN pools from the week ending on 7 March. These data indicate that SARS-CoV-2 infections were present in a small number of patients seeking care at MSHS facilities across NYC several weeks prior to the first pandemic wave. The high number of positive RPN pools in the first week of March provides an explanation for the “sudden” exponential increase in severe COVID-19 cases that were admitted to MSHS hospitals starting mid-March 2020.

**Fig. 1.**
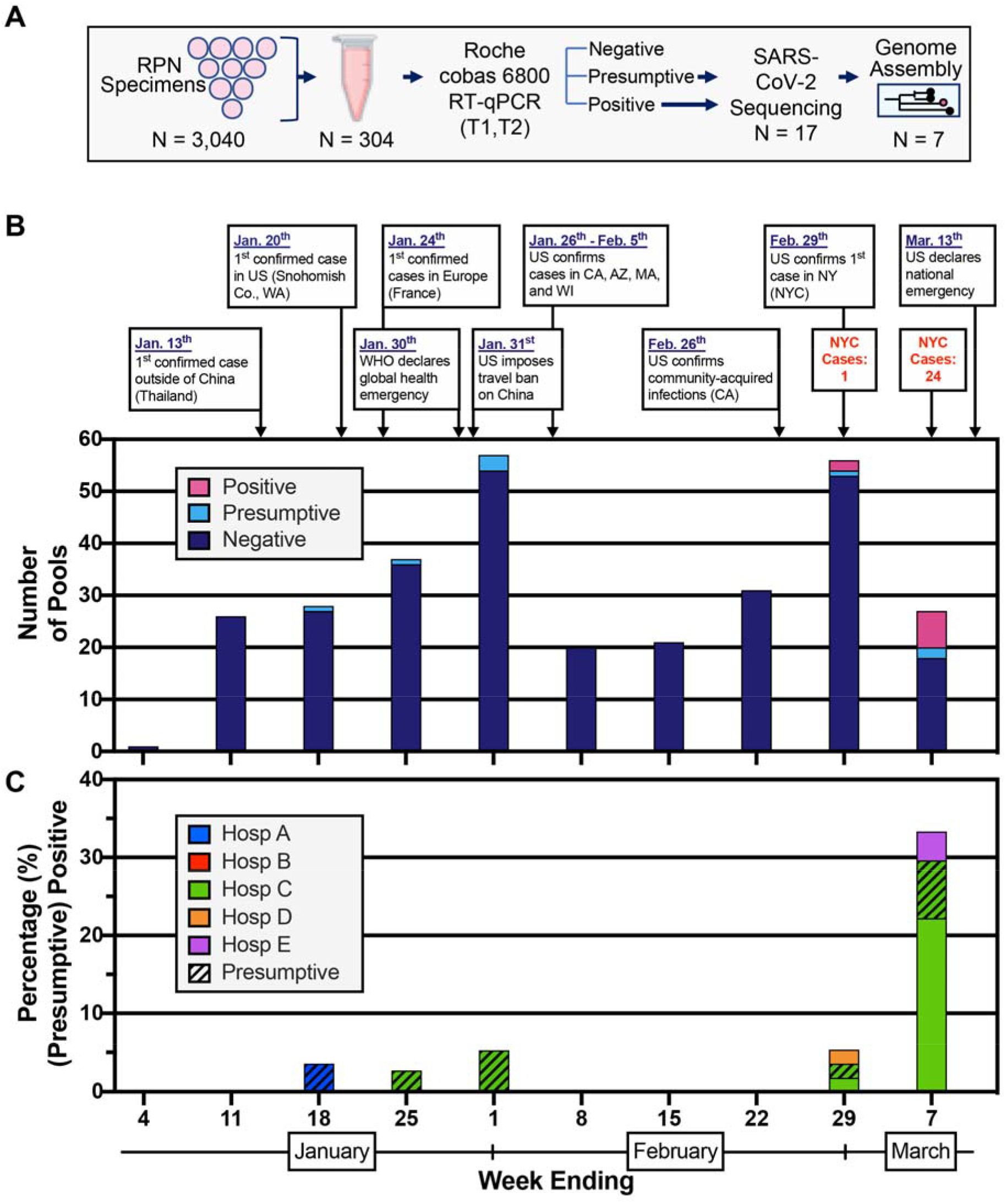
Detection of SARS-CoV-2 nucleic acids in nasopharyngeal specimens collected in the first ten weeks of 2020. (A) Schematic representation of the study design. Nasopharyngeal swab specimens that tested negative for respiratory pathogens (RPN) were pooled. Each pool consisted of 10 specimens from the same week from one of five hospital sites. Nucleic acid amplification testing (NAAT) was performed and RNA was processed for SARS-CoV-2 genome assembly. (B) Select events and responses to the evolving SARS-CoV-2 pandemic are annotated over the timeframe surveyed. Confirmed cases in NYC for the last two weeks are noted. Absolute counts of pools that tested positive, presumptive positive, and negative by RT-PCR are depicted by week collected. (C) Distribution of positive (solid) and presumptive positive (hatched) pools across the five different hospital sites in NYC.

To validate the NAAT results and to reconstruct the SARS-CoV-2 genomes in these pooled RPN specimens, we extracted viral RNA from all positive and presumptive positive RPN pools and performed viral genome sequencing as described previously (*4*). We obtained complete SARS-CoV-2 genomes with distinct genotypes from six of the nine positive pools (**Fig. 2A**). To assess for the presence of more than one distinct viral genome in these pools, we determined the fraction of non-consensus viral variants for all positions in each assembly. The maximum fraction of non-consensus variants at any position did not exceed 20%, suggesting that each pool was dominated by a single viral variant.

**Fig. 2.**
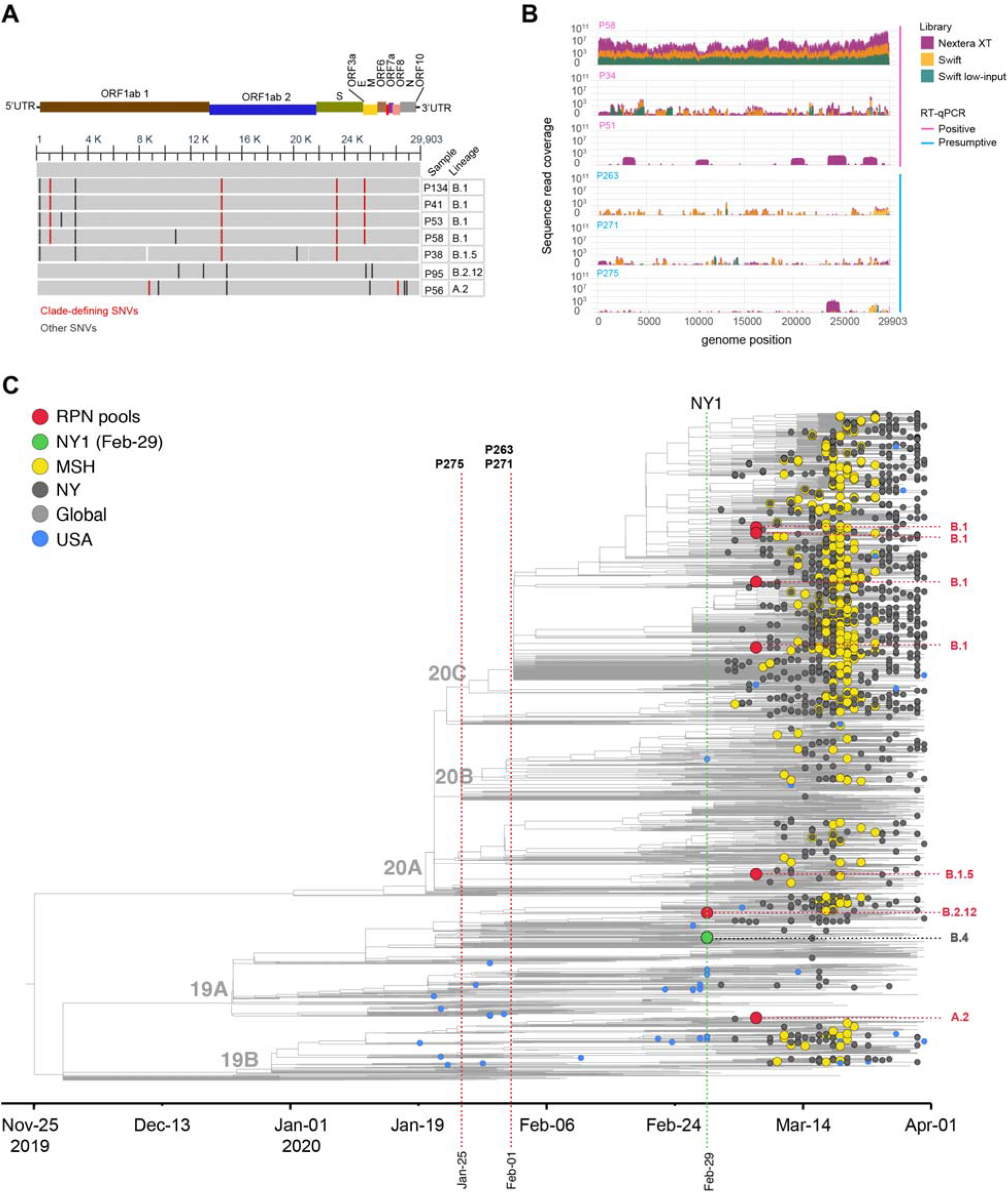
Phylogenetic relationships of previously undetected SARS-CoV-2 and other NY and global isolates. (A) Multiple sequence alignment of SARS-CoV-2 genome sequences obtained from RPN pools containing specimens collected prior to the first confirmed case in NY (NY1) relative to Wuhan-Hu-1 (RefSeq: NC_045512). RPN pools are ordered by date and lineage as displayed in panel A. The SARS-CoV-2 genome coordinates and gene annotations are shown above. Single nucleotide variations (SNVs) are depicted with vertical lines in red (clade-defining) or black (other). (B) Sequence read coverage for three sequencing libraries of partially sequenced RT-qPCR positive (magenta) and presumptive (cyan) specimens with detectable SARS-CoV-2 reads >Q30 reads are shown. (C) Maximum likelihood (ML) phylodynamic inference of seven SARS-CoV-2 genome sequences from this surveillance study in a global background of 2,993. Tip circles indicate the position of the respiratory pathogen-negative (RPN) pools (red) described in this report, the first reported COVID-19 case in New York City (green) from February 29, later NYC cases from MSHS (yellow) and other institutions (dark grey), and global (light grey) and US (blue) early isolates prior to 1 March. The PANGOLIN lineage classification of the RPN pools is indicated on the right, and the NextStrain clades are shown as node labels. The specimen identifier is indicated for RPN pools detected earlier than NY1. The time tree was inferred under a strict clock model with a nucleotide substitution rate of 0.80×10^−3^.

Three NAAT positive and eight NAAT presumptive positive RPN pools yielded either scattered or no SARS-CoV-2 reads, suggesting that viral RNA levels in these pools were insufficient to obtain complete genomes. Indeed, these positive pools had high Ct values for both targets by NAAT assay (e.g., ≥34.25 for T1 (Orf1ab), ≥35.63 for T2 (E)) (**Fig. S1B**). We performed a second viral genome sequencing protocol with smaller tiling amplicons (see Methods) to improve recovery of degraded viral RNA. These additional sequencing data allowed us to complete another SARS-CoV-2 genome from positive pool P58 (**Fig. 2A**). The remaining two positive pools yielded partial genomes (35% genome completeness for P34, and 24% for P51) (**Fig. 2B**). We were not able to assemble consensus genome sequences from any of the presumptive positive samples, but three pools from weeks ending 25 January (P275) and 1 February (P263 and P271) each yielded scattered SARS-CoV-2 reads throughout the viral genome (**Fig. 2B, Table S1**), confirming the presence of viral genetic material. Clade-defining sites were not sufficiently covered to assign these pools to specific clades or lineages.

We next reconstructed phylogenetic relationships between each of the seven early complete genomes (≥95% genome coverage) and a representative dataset of available genomes from the US as well as from viruses circulating globally between January and March 2020 (**Fig. 2C**). In order to place these genomes on a timed tree reconstruction, we conservatively used the week ending date of each pool. All RPN SARS-CoV-2 genomes were identified in specimens collected in the last week of February (ending 29 February) and the first week of March (ending 7 March), a time period when molecular diagnostic testing still was limited to individuals fulfilling a very narrow range of testing criteria. The sequences from these early infections map to four different lineages, B.1 (n=4), B.1.5 (n=1), B.2.12 (n=1), and A.2 (n=1), consistent with multiple independent introductions (Fig. 2C, **Table 1**). All four lineages were detected subsequently during the peak in the spring of 2020 in NYC, which was dominated by the B.1 lineage (*4, 5*). Notably, the B.1 RPN pools (P134, P41, P58, P53) are nested within a cluster that was linked to early community spread of SARS-CoV-2 in NYC, delineated by the additional substitutions ORF3a-Q57H and ORF1a-T265I (*4, 5*).

**Table 1.** Lineage classification of previously undetected SARS-CoV-2 in NYC.

| Sample | Week-ending | Genome completeness (%) | NextStrain Clade | Clade-defining mutations | PANGOLIN Lineage | Lineage detection prior to 1 March |
| --- | --- | --- | --- | --- | --- | --- |
| Pool-134 | 7-Mar-2020 | 99.9 | 20C | S:D614G,<br>ORF1b:P31<br>4L,ORF3a:<br>Q57H,<br>ORF1a:T26<br>5I | B.1 | Mainly Europe, linked to Italian outbreak, only a few North American (non-US) isolates |
| Pool-41 | 7-Mar-2020 | 99.7 | 20C | S:D614G,<br>ORF1b:P31<br>4L,ORF3a:<br>Q57H,<br>ORF1a:T26<br>5I | B.1 |  |
| Pool-58 | 7-Mar-2020 | 99.8 | 20C | S:D614G,<br>ORF1b:P31<br>4L,ORF3a:<br>Q57H,<br>ORF1a:T26<br>5I | B.1 |  |
| Pool-53 | 7-Mar-2020 | 99.8 | 20C | S:D614G,<br>ORF1b:P31<br>4L,ORF3a:<br>Q57H,<br>ORF1a:T26<br>5I | B.1 |  |
| Pool-38 | 7-Mar-2020 | 98.9 | 20A | S:D614G,<br>ORF1b:P31<br>4L | B.1.5 | Europe/South America/Asia |
| Pool-95 | 29-Feb-2020 | 99.8 | 19A | T14408C | B.2.12 | Asia/Europe/Oceania |
| Pool-56 | 7-Mar-2020 | 99.7 | 19B | C8782T<br>ORF8:L84S | A.2 | Europe |

Our study has several limitations, because the RPN pools were made from available residual diagnostic specimens that varied with respect to duration and conditions of storage. It is, therefore, possible that some positive specimens - particularly those with low viral titers - were missed due to degradation of the viral RNA genomes. We started systematically banking RPN specimens in February 2020 and, as a result, may have missed some RPN specimens obtained in early January. However, we included all available residual RPN specimens in our study without any selection. Furthermore, although we reconstructed a single, dominant viral genome from each pool, it is possible that other distinct SARS-CoV-2 variants were present at lower levels. Thus, our estimates regarding the frequency of SARS-CoV-2 positivity over time are a conservative approximation. Lastly, we lack demographic and epidemiological information for individual cases since we relied on de-identified pooled specimens.

Taken together, we provide clear evidence that SARS-CoV-2 infections were present in NYC at least 6 to 8 weeks prior to the surge of cases that flooded the NYC health system. Previous studies have suggested cryptic transmissions weeks prior to the first confirmed cases of community spread (*8, 9*). Large retrospective testing efforts have probed for SARS-CoV-2 in banked nasopharyngeal specimens from at least seven states (Michigan, Pennsylvania, Tennessee, Texas, Wisconsin, Washington State (*10*), and California (*11*)), with the earliest positive specimens dating back to 21 February 2020 (Seattle, WA (*9*) and California (*11*)). In addition, a recent serosurvey of blood products further suggests early undetected spread in multiple states across the US from December 2019 through January 2020 (*12*). Of note these studies relied solely on molecular testing without validation by viral genome sequencing. Our study is complementary to those efforts, and provides information regarding the presence of SARS-CoV-2 in the diverse, densely populated, international travel hub of NYC, more than one month prior to the detection of the first reported NYS case.

Although we detected SARS-CoV-2 RNA in specimens from late January 2020, without fully reconstructed genomes it is impossible to determine whether these cases seeded the community spread observed later in March. Our molecular findings are in agreement with previous evidence of sporadic SARS-CoV-2 infections in the US in January 2020 (9), and are consistent with evidence from a MSHS SARS-CoV-2 serosurvey (*6*) that identified low levels of SARS-CoV-2 antibody positivity as early as mid-February 2020. Lastly, although our survey only examined RPN specimens collected starting 30 December 2019, the absence of SARS-CoV-2 positive specimens from early January, in conjunction with the serological evidence (*6*), makes the presence of SARS-CoV-2 in the US East Coast populace prior to 2020 unlikely.

The observation that the majority of SARS-CoV-2 genomes identified from RPN pools including specimens collected between the last week of February and the first week of March 2020 cluster within the B.1 lineage is consistent with phylogenetic analyses by us and others linking most cases during the first wave to an influx of travelers from Europe (*4, 5, 7*) prior to travel restrictions on mainland European countries (on 13 March 2020) and the United Kingdom and Ireland (on 16 March 2020). Our findings provide further evidence that the limited availability of diagnostic testing early in the epidemic hindered the identification of SARS-CoV-2-infected individuals (*8, 10, 13, 14*) and help explain the expansion of the epidemic notwithstanding travel restrictions designed to limit further introductions of SARS-CoV-2 into the US. These observations indicate a brief window of opportunity in which surveillance, testing, and contact-tracing of a limited number of infections may have stemmed community spread.

Systematic, unbiased surveillance of clinical specimens obtained from individuals presenting with unexplained or unusual clinical presentations of respiratory illness for the presence of emerging viral pathogens must be a key component of any future early-warning sentinel programs. Population-dense metropolitan areas and major global travel hubs present not only a heightened risk for community spread but also an opportunity for monitoring and prevention. These systematic measures will need to become essential components of our new normal in order to prevent local infections and transmissions from blooming into uncontrolled outbreaks.

## MATERIALS AND METHODS

### Ethics statement

This study was reviewed and approved by the Institutional Review Board of the Icahn School of Medicine at Mount Sinai (protocol: HS# 20-00141).

### SARS-CoV-2 specimen collection and testing

Respiratory pathogen-negative (RPN) pools in this study were compared to later sequences obtained from individual clinical specimens from cases that tested positive for SARS-CoV-2 in the Mount Sinai Health System (MSHS) once testing became more widely available. Details on testing using the aforementioned systems were previously described (4).

### Preparation of respiratory pathogen-negative (RPN) pools

RPN pools were prepared by mixing aliquots from nasopharyngeal specimens in viral transport medium from patients with respiratory symptoms that previously tested negative for routine respiratory pathogens using multiplex diagnostic panels (*e*.*g*., BioMerieux FilmArray Respiratory Panel, Cepheid Xpert® Xpress Flu/RSV). RPN specimens (n=3,040) collected at MSHS hospitals and outpatient facilities between 30 December 2019 and 7 March 2020 were organized into groups of ten, and stored at −80°C. Notably, these specimens had not previously been tested for SARS-CoV-2. The RPN pools (n=304) were prepared in an isolated class II biological safety cabinet at a separate location from the Clinical Microbiology and research labs, that had never been used for handling respiratory specimens or viruses.

Briefly, 400µL of viral transport medium from each specimen was manually aliquoted one-at-a-time into a sterile 5 mL snap-cap centrifuge tube (ASi, C2520). Once each specimen was aliquoted, the 4 mL volume was mixed manually by pipetting and 600 μL aliquots were reserved for SARS-CoV-2 nucleic acid amplification testing (NAAT). RPN specimens and pools were stored at −80°C.

### SARS-CoV-2 nucleic acid amplification testing (NAAT)

To test for SARS-CoV-2 in RPN pools, 600 μL aliquots underwent NAAT by the cobas® 6800/8800 SARS-CoV-2 real-time RT-PCR Test (Roche, 09175431190) in the MSHS Clinical Microbiology Laboratory, which is certified under Clinical Laboratory Improvement Amendments of 1988 (CLIA), 42 U.S.C. §263a and meets requirements to perform high complexity tests. Aliquots were run in batches with one cobas® Buffer Negative Control (BUF (-) C) (Roche, 07002238190) and one cobas® Positive Control (SARS-CoV-2 (+)C) (Roche, 09175440190). The assay utilizes two targets to detect SARS-CoV-2 RNA: the SARS-CoV-2-specific Orf1ab gene (T1) and the pan-Sarbecovirus envelope E gene (T2). All target results were valid across all 304 RPN pools tested. A result was deemed positive for SARS-CoV-2 if both T1 and T2 were detected, or if T1 was detected alone. A result was deemed presumptively positive if T2 was detected alone. A result was deemed negative if neither T1 nor T2 was detected

### Optimized extraction of total RNA from pools

Total RNA was extracted manually from 1mL aliquots of each positive or presumptively positive pool, utilizing the QIAamp UltraSens Virus Kit (QIAGEN, 53706) and using an optimized protocol. Prior to extraction, all RPN pools were equilibrated to room temperature for at least thirty minutes. Briefly, 1mL of the pooled viral transport medium was transferred to a 2mL Dolphin Tube (Genesee Scientific, 24-284), lysed by adding 800μL Buffer AC, and manually mixed by pipetting up and down. Carrier RNA (5.6 μL) was added to each tube and each mixture was vortexed one-at-a-time. Lysates were incubated at room temperature for 10 minutes and spun at 5,000 g for 3 minutes. Tubes were opened and supernatants removed from each tube within a biological safety cabinet,.

Lysates were moved to an isolated clean research space designated for nucleic acid extraction. A mixture of Buffer AR (300 μL) and proteinase K (20 μL) pre-warmed to 60°C was added to each lysate which was then vortexed for 20 seconds. Lysates were incubated on a ThermoMixer C (Eppendorf, 2231000667) at 40°C, shaking at 2,200 rpm for 10 minutes. Lysates were then spun down and 300μL Buffer AB was added to each tube. Mixtures were vortexed for 10 seconds and RNA was purified by manual extraction on QIAamp spin columns and eluted in 50μL of AE Elution Buffer for downstream confirmatory RT-PCR testing and sequencing applications.

### SARS-CoV-2 whole-genome amplification and sequencing

All positive and presumptive positive pools were sequenced on the Illumina MiSeq platform following ProtoScript II (New England Biolabs, E6560) cDNA synthesis with random hexamers, SARS-CoV-2 whole-genome amplification with custom designed tiling primers, and library preparation with the Nextera XT DNA Sample Preparation kit (Illumina, FC-131-1096), as described previously (4).

For each pool that did not yield a complete genome in the initial sequencing attempt, 4 additional sequencing libraries were prepared from re-extracted RNA. 1) Nextera XT Illumina amplicon sequencing as described above, 2) Nextera XT sequencing of 1.5 to 2kb amplicons targeting only regions containing clade-defining SNVs (positions 1059, 8782, 14408, 23403, 25563, 28144, 28881 and 28882, https://nextstrain.org/blog/2020-06-02-SARSCoV2-clade-naming), and the Swift Normalase® Amplicon Panel (SNAP) SARS-CoV-2 (Swift Bioscience COVG1V2-96, SN-5×296 and SN-5S1A96) according to the manufacturer’s instructions for 3) regular input and 4) low input samples. Data from the Nextera XT libraries were combined for assembly.

### SARS-CoV-2 genome assembly

Illumina data were analyzed using a custom reference-based (MN908947.3) pipeline (15), https://github.com/mjsull/COVID_pipe, to reconstruct SARS-CoV-2 genomes, as previously described (4).

### SARS-CoV-2 phylogenetic analysis and lineage assignment

Phylogenetic relationships of the 7 high-quality consensus sequences (>80% completeness) were inferred over a global background of SARS-CoV-2 sequences between December 2019 and May 2020 downloaded from GISAID as previously described (4) with a few modifications. For the background set, only sequences with >5% non-ambiguous sites were included, and sequences were masked at the 5’ and 3’ ends to remove ambiguous regions but conserve UTR regions that contained SNVs across the whole data set. Initial alignment and subsampling were done by using the NextStrain tool (16). For cases with available information on epidemiological links, or patients with longitudinal sampling when known, only one representative sequence was kept. A maximum likelihood (ML) phylogeny was inferred under the GTR+F+I+G4 model (17, 18), after which further manual curation was done to identify and remove extreme outliers that deviated from a temporal signal using Tempest (19). The final ML tree was then time-scaled with TreeTime using the strict and relaxed clock models as previously described (4).

Lineage classification was done using a phylogenetic based nomenclature as described by Rambaut *et al*. (20) using the PANGOLIN tool, lineages version 2020-10-03 (21).

## Data Availability

SARS-CoV-2 genome consensus sequences for all study isolates and sample pools were deposited in GISAID.

## ACKNOWLEDGEMENTS

We thank all the members of the Simon and van Bakel laboratories for pitching in whenever additional help was needed. We also acknowledge the invaluable help and continuous assistance provided by Rapid Response Laboratories and Clinical Microbiology Laboratory of the Mount Sinai Health System with regard to the banking and transfer of nasopharyngeal specimens. We thank Denis Ruchnewitz and Michael Lässig for their input on the phylogenetic analyses as well as Catherine Teo for her efforts in RT-qPCR assays. We are grateful for the continuous expert guidance provided by the ISMMS Program for the Protection of Human Subjects (PPHS). We also acknowledge the authors and the originating and submitting laboratories of sequences from GISAID’s EpiFlu and EpiCoV (www.gisaid.org) that were used as background for our phylogenetic inferences.

## SUPPLEMENTARY MATERIALS

**Fig S1.**
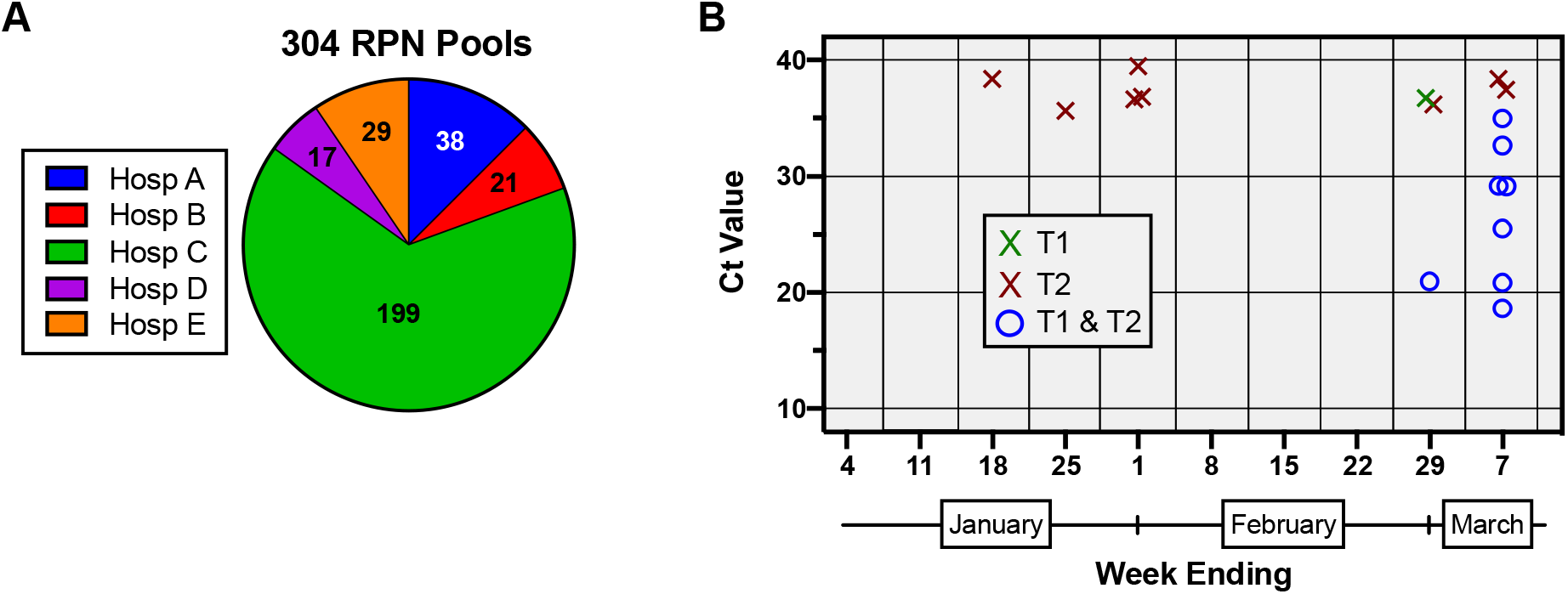
Hospital distribution and SARS-CoV-2 quantitation of negative pools. (A) 304 RPN pools were generated from nasopharyngeal swab specimens collected at distinct MSHS sites (A-E) from 30 December 2019 to 7 March 2020. (B) SARS-CoV-2 NAAT quantities in RPN pools by week. Cycle thresholds (Ct) of pools yielding a positive or presumptive positive result by clinical diagnostic SARS-CoV-2 RT-PCR are depicted. If only assay target 1 (T1, ORF1ab) or only assay target 2 (T2, E-gene) was detected in a pool, an X denotes the corresponding Ct value. If both T1 and T2 were detected, the average of the Ct values of both detected targets is depicted by a circle for that pool.

**Table S1.**
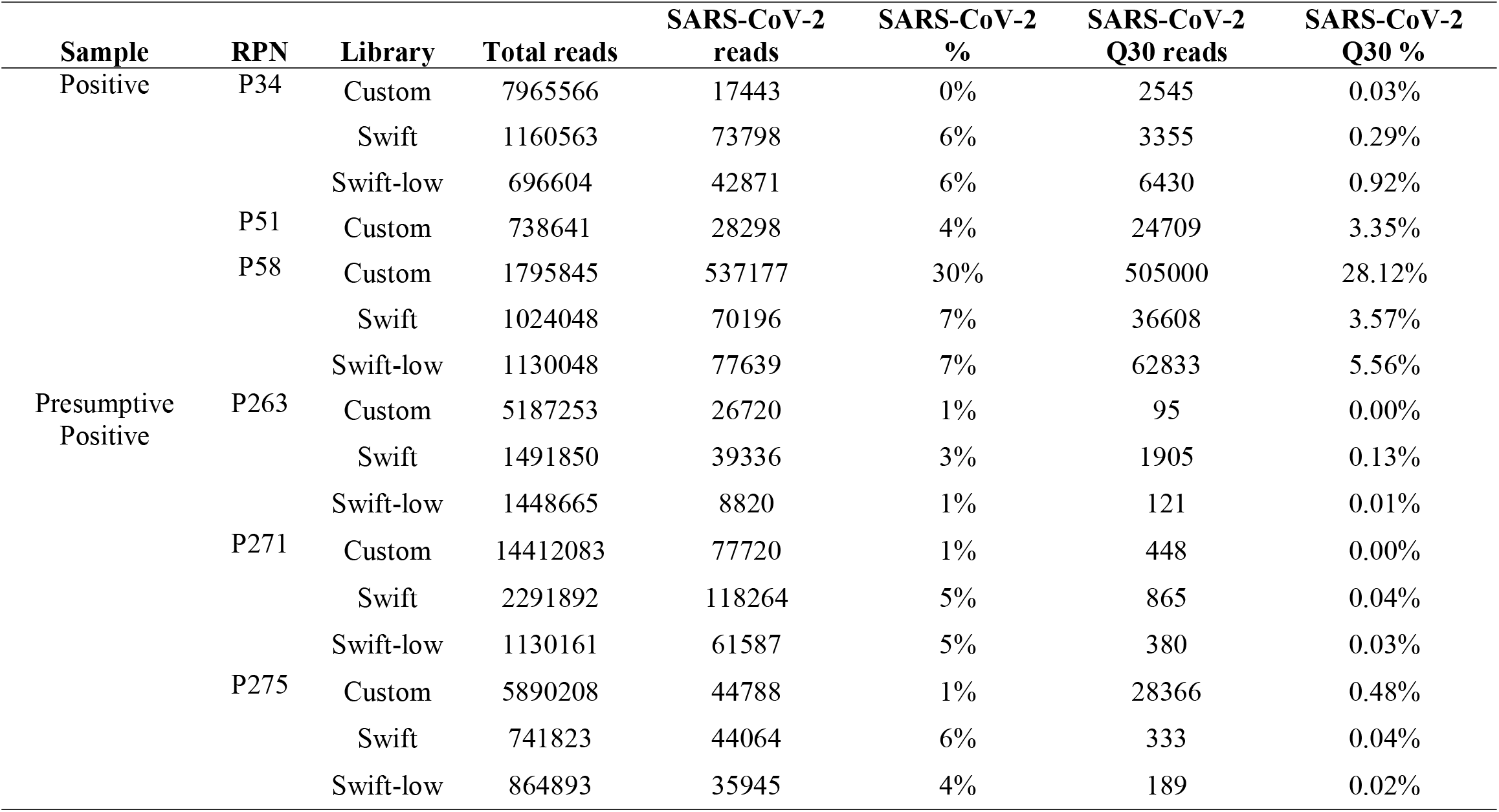
Total and SARS-CoV-2 mapped reads per library for RNPs with incomplete genomes.

